# Escalating combinations of enhanced infectivity and immune escape define SARS-CoV-2 Omicron lineage replacement

**DOI:** 10.1101/2024.01.03.24300790

**Authors:** Nicholas F.G. Chen, Kien Pham, Chrispin Chaguza, Rafael Lopes, Fayette Klaassen, Daniel M. Weinberger, Virginia E. Pitzer, Joshua L. Warren, Nathan D. Grubaugh, Anne M. Hahn

## Abstract

In 2022, consecutive sweeps of the highly transmissible SARS-CoV-2 Omicron-family maintained high viral transmission levels despite extensive antigen exposure on the population level resulting from both vaccinations and infections. To better understand variant fitness in the context of the highly dynamic immunity landscape of 2022, we aimed to dissect the interplay between immunity and fitness advantages of emerging SARS-CoV-2 Omicron lineages on the population-level. We evaluated the relative contribution of higher intrinsic transmissibility or immune escape on the fitness of emerging lineages by analyzing data collected through our local genomic surveillance program from Connecticut, USA. We compared growth rates, estimated infections, effective reproductive rates, average viral copy numbers, and likelihood for causing vaccine break-through infections. Using these population-level data, we find that newly emerging Omicron lineages reach dominance through a specific combination of enhanced intrinsic transmissibility and immune escape that varies over time depending on the state of the host-population. Using similar frameworks that integrate whole genome sequencing together with clinical, laboratory, and epidemiological data can advance our knowledge on host-pathogen dynamics in the post-emergence phase that can be applied to other communicable diseases beyond SARS-CoV-2.

## Introduction

Capacities for genomic surveillance saw an unprecedented rise during the COVID-19 pandemic (Brito et al. 2022). The ability to closely monitor SARS-CoV-2 populations in real-time was crucial to discover and track SARS-CoV-2 variants of concern (VOC). The first VOC, Alpha (B.1.1.7) (Hill et al. 2022; Rambaut et al. 2020), emerged towards the end of 2020 and was followed in 2021 by Beta (B.1.351) (Tegally et al. 2021), Gamma (P.1) (Faria et al. 2021), and Delta (B.1.617) (WHO 2021). In late 2021, the highly divergent Omicron (B.1.1.529) lineage (Viana et al. 2022) rapidly displaced Delta globally.

In 2022, SARS-CoV-2 transmission remained, somewhat counterintuitively, high all year, which stands in contrast to the summer-lows observed during 2020 and 2021. This was characterized by consecutive sweeps of multiple Omicron lineages. BA.1 (B.1.1.529.1), the first member of the Omicron family to emerge, shared a common ancestor with the D614G background in late 2020. This ancestor diverged into at least five main sublineages (BA.1-BA.5) (Tegally et al. 2022; Viana et al. 2022). The exact origin and nature of the emergence of these Omicron lineages remains unclear; however, it is likely that these lineages emerged from a common source, likely undergoing selective pressure for an extended amount of time (Markov et al. 2023). A subset of the original Omicron viral population further spread in one or multiple transmission events to a larger community and eventually worldwide (Tegally et al. 2023).

Thus, 2022 saw a unique combination of this highly divergent virus variant facing a host population that, through intensive vaccination campaigns and vaccine roll-out in many parts of the world, had built up significant amounts of immunity towards the original Wuhan-Hu-1 Spike antigen. In this study, we aimed to reconstruct key epidemiological and evolutionary features that led to the particular lineage replacement dynamics of Omicron lineages as exemplified in a well-monitored study setting in Connecticut, United States (US). For this, we drew upon sample data collected from the Yale New Haven Hospital (YNHH) through our Yale Genomic Surveillance Initiative from January 2022 to January 2023. Whole genome sequencing data together with relevant laboratory and patient metadata were available for approximately 5-10% of total reported cases across the state and the entire length of the study period. To characterize competitive growth advantages of variants during their emergence periods and evaluate their fitness compared to their immediate predecessors, we calculated the growth rates of each incoming Omicron lineage, compared their virus copy number from nasal swabs, and calculated the risk for vaccine breakthrough infections as a metric for immune escape. Doing so, we reveal a high capacity of Omicron lineages to explore different fitness niches through a combination of enhanced transmissibility and/or immune escape depending on the history of antigenic exposure of the host-population.

The synthesis of high-resolution epidemiological, genomic, and immunological data allows us to assess and compare transmissibility and immune escape of each Omicron lineage. Exploring lineage replacement dynamics on a population level allows us to evaluate predictions based on *in-vitro* data and thus evaluate key parameters defining future variant emergence. Doing so, we reveal a high capacity of Omicron lineages to explore different fitness niches through a combination of enhanced transmissibility and/or immune escape depending on the history of antigenic exposure of the host-population. Our study highlights the ability to derive key aspects of pathogen lineage fitness by analyzing data that can be drawn from genomic surveillance efforts with relatively simple technical requirements. Such frameworks will be particularly relevant for further monitoring SARS-CoV-2 in the post-pandemic phase and are easily transferable to other pathogens.

## Results

### Continuous Omicron lineages replacement causes high levels of community transmission throughout 2022

Close monitoring of the SARS-CoV-2 population composition added valuable insights on how emerging variants were impacting COVID-19 case dynamics. The first VOCs detected in Connecticut were Alpha in late 2020 and Delta around mid-2021 (**Fig 1A**, based on data from GISAID for Connecticut). Looking at the epidemic curve in Connecticut (**Fig 1B**), the introduction of both the Alpha and Delta VOCs were followed by a surge in reported SARS-CoV-2 cases in December 2020 and Spring 2021, respectively. After a summer of relatively low transmission in 2021, cases started to rise again towards the end of the year with a second Delta-dominated peak in December 2021.

**Fig 1.**
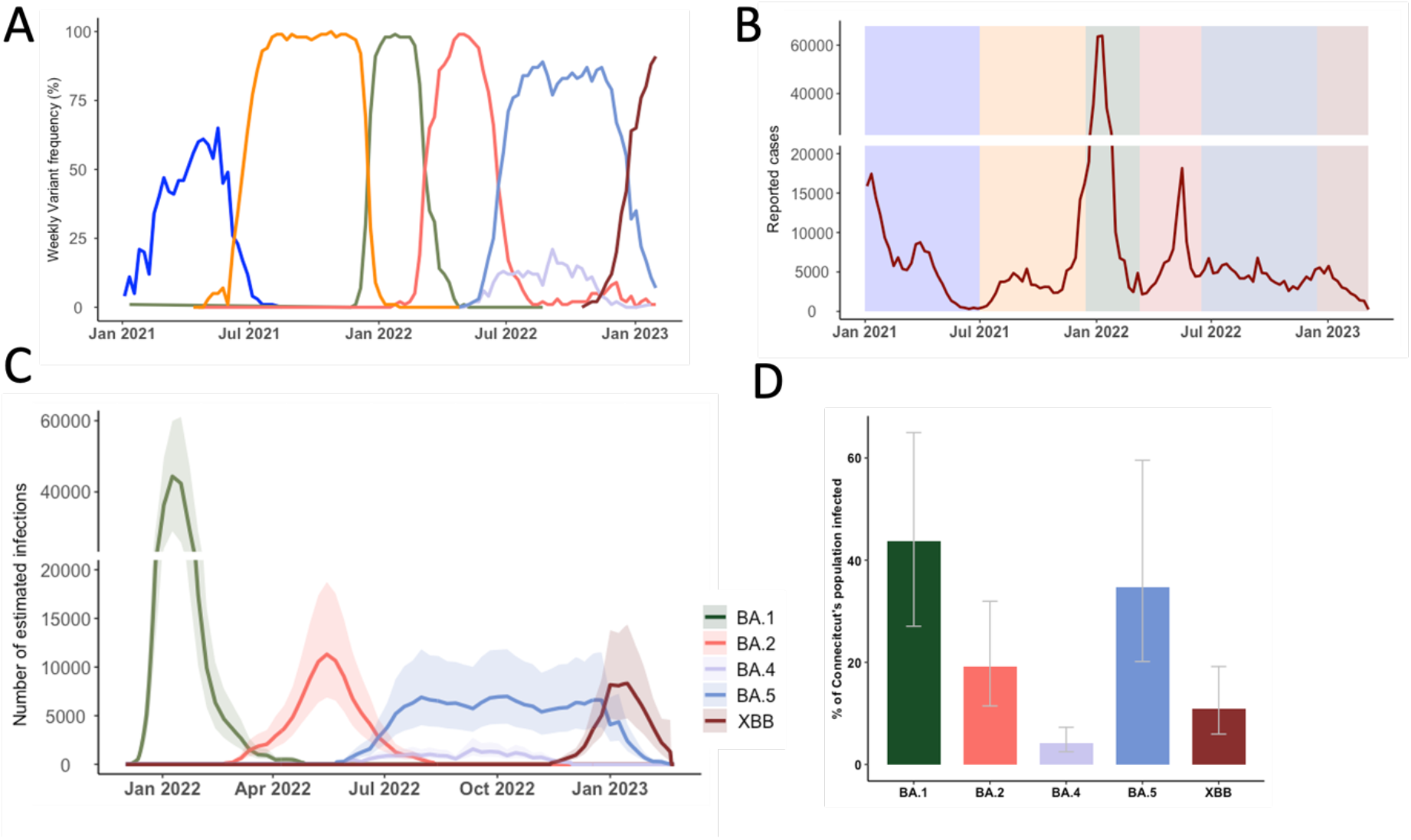
Genomic Epidemiology of SARS-CoV-2 describes the COVID-19 epidemic in Connecticut, US. **(A)** Frequencies of SARS-CoV-2 variants of concern from January 2021 to January 2023, based on sequences deposited on GISAID. **(B)** Reported COVID-19 infections from January 2021 to 2023 with highlights of the dominant periods of different Omicron lineages, data from the Connecticut Department of Public Health **(C)** Number of estimated infections based on the covidestim model for 2022 where testing and reporting widely changed after the first wave of Omicron BA.1 **(D)** Cumulative estimated cases for each Omicron variant up until January 2023.

Globally, the first Omicron sample was identified in November 2021 and shortly after also detected in the US and Connecticut (**Table 1**). BA.1 is an outgroup to the BA.2/BA.4/BA.5 cluster, and XBB a recombinant of two second generation BA.2 lineages (BA.2.10 & BA.2.75) that likely arose in Southeast-Asia around late summer 2022 (Goh et al. 2023) (**Supplementary Fig 1**). Notably, whereas BA.1-BA.5 most likely originated from a common source (Tegally et al. 2021, 2022; Viana et al. 2022), XBB derived from lineages that emerged through sustained transmission chains in 2022. After BA.1’s emergence in Connecticut in November 2021, the rapid replacement of Delta was accompanied by a massive increase in reported cases. Then, BA.2 swiftly replaced BA.1 in March 2022. BA.4 and BA.5 were both introduced to Connecticut in May 2022 and co-circulated for eight months until December 2022 (**Table 1**). Lastly, XBB-based variants outcompeted BA.5 lineages towards the end of 2022. Interestingly, the New England region including our study site was the first where XBB.1.5 was widely circulated within the US (Ma et al. 2023).

**Table 1.** Major Omicron lineages in Connecticut January 2022 - January 2023, info taken from GISAID.

| Lineage | First detected globally | First detected in CT | Major sub-lineages in Connecticut (+key amino acid changes in Spike) |
| --- | --- | --- | --- |
| BA.1 | October 2021 | November 2021 | BA.1.1 (+R346K) |
| BA.2 | November 2021 | January 2022 | BA.2.12.1 (+L452Q,S704L) |
| BA.4 | November 2021 | May 2022 | BA.4.6 (+R346T, N658S) |
| BA.5 | November 2021 | May 2022 | BQ.1.1 (+R346T, K444T,N460K)<br>BF.7 (+R346T) |
| XBB (BA.2.10*x<br>BA.2.75*) | September 2022 | October 2022 | XBB.1.5 (+F486P) |

To understand the factors that influenced the consecutive sweeps of Omicron lineages, we first characterized the infection dynamics broken up for each lineage with respect to case counts across our one-year study period. As the northern hemisphere entered into the summer months of 2022, most pandemic response mechanisms and non-pharmaceutical measures were discontinued. Consequently, case reporting is expected to be less reliable in accurately mirroring true infection dynamics compared to earlier in the pandemic. Therefore, we retrieved data from covidestim, a Bayesian nowcasting model incorporating publicly available time series of COVID-19 case notifications, hospitalizations, and deaths, accounting for effective population immunity to estimate infections (Chitwood et al. 2022). Together with the frequencies of the major Omicron lineages derived from state-wide genomic surveillance data, we partitioned the overall estimated infections to calculate variant-specific infection estimates cumulatively and over time.

By analyzing the variant-specific estimated infections generated through covidestim, we are further able to characterize dynamics of each lineage that may not be reflected in reported cases **(Fig 1C).** Using this framework, we estimated that the BA.1 lineage caused approximately 1.5 million infections in Connecticut, USA (**Fig 1D**), representing around 44% (Credible Interval (CrI) 27%-65%) of the state’s population. Interestingly, the BA.2, BA.4, and BA.5 lineages caused over 2.1 million combined infections (CrI 1.2-3.5 million) within only nine months after this surge (**Fig 1D**). Another surge driven by the XBB lineage and its descendants caused an estimated 0.4 million infections (CrI 0.2-0.7 million) up until January 2023.

Taken together, we reconstructed in detail the SARS-CoV-2 infection dynamics caused by Omicron sublineages in 2022 by drawing upon data from genomic surveillance and epidemiological modeling. We showed the timing and scale of each of the genetically closely related Omicron lineages differed substantially and we thus aimed to further characterize each of the emergence periods.

### Growth advantage of emerging Omicron lineages shrinks towards the end of 2022

Having established that each of these variants exhibits distinct epidemic dynamics in the population, we aimed to further explore the specific features of each of the variants and how these contributed to this emergence pattern. For this, we first determined the rate of lineage replacement by calculating the time it took for each lineage to increase from 5% to 50% of all reported daily cases in Connecticut (Fig 2A). This range of 5% to 50% was chosen to capture the period of time it took for a variant to become reliably established in the population, while avoiding some of the inherent stochasticity associated with initial cases, to when it accounted for the majority of cases and is thus dominant in the population. We then fitted a logistic regression model to the observed frequencies and compared their slope coefficients **(Fig 2B)**.

**Fig 2.**
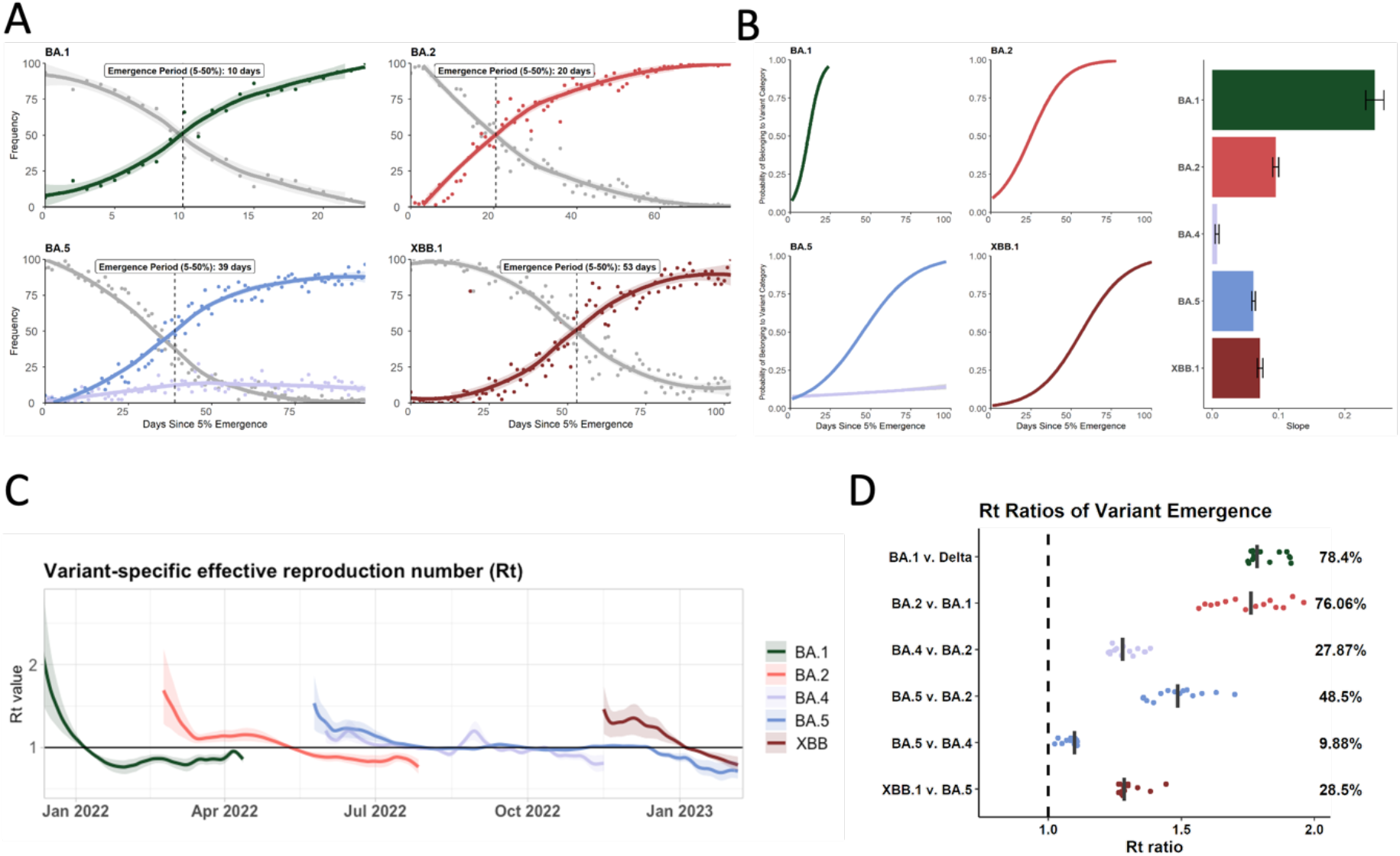
Comparison of key epidemiological parameters for each Omicron lineage in 2022. **(A)** Frequencies of the incoming Omicron lineage during its emergence period highlight the time interval between being detected at 5% and 50% of all sequences (based on GISAID data for Connecticut). **(B)** Comparison of growth rates between the different Omicron lineages and the average slope of the growth curve during emergence periods **(C)** Variant-specific R_t_ numbers for each of the Omicron lineage over time derived from the modeled overall R_t_ from the covidestim model **(D)** Based on (C), the R_t_ ratios for each of the emergence periods were calculated to estimate the average advantage of each incoming lineage compared to the previously dominating one.

Doing so, we show that BA.1 had the fastest growth rate, reaching a frequency of 50% in only 10 days from being detected at 5% **(Fig 2A)**. For each of the subsequent lineages, lineage replacement slowed down, ranging from 20 to 59 days for BA.2 and XBB, respectively.

We further compared the emergence of each of these lineages using variant-specific estimated infections from the covidestim model. For this, we partitioned the covidestim weekly estimated COVID-19 infections in Connecticut based on state-wide variant frequency to calculate variant-specific reproduction numbers (R_t_) over the course of 2022 (**Fig 2C**). We show that although BA.1 had the highest initial R_t_ value (2.4, CrI 1.6-3.5), it quickly fell below 1 in early January. BA.2 emerged in February 2022 with an initial R_t_ of 1.7 (CrI 1.3-2.2), followed by a notable decrease and a plateau at around 1.2 for several weeks before falling below 1 around April 2022. The initial R_t_ of BA.5 (1.5 CrI 1.2-1.86) and BA.4 (1.2 CrI 1.07-1.36) were overall lower than for BA.1 and BA.2. However, BA.4 and BA.5 both hovered at R_t_ ∼ 1 for several months during summer 2022. BA.4 was finally outcompeted by late-stage BA.5 lineages. The R_t_ for BA.5 fell below 1 once XBB was introduced with an R_t_ of 1.4 (CrI 1.1-1.7). The R_t_ of XBB fell below 1 around the beginning of 2023.

Next, we sought to understand the relative transmission advantages during the emergence periods by comparing R_t_ values of co-circulating lineages (**Fig 2D**). We first identified 14-day windows in which one lineage replaced the previously dominating lineage (**Supplementary Table 1**). Using a method previously developed (Earnest et al. 2022; Petrone et al. 2022), we identified six, 14-day periods of variant overlap and divided the R_t_ value of the emerging lineage by the R_t_ value of the previously dominating lineage for each day of the emergence period, yielding 14 R_t_ ratios. The median of these values results in a comparative fitness measure (**Fig 2D**). We show that BA.1 and BA.2 were 78.4% and 76% more transmissible than Delta and BA.1, respectively (**Fig 2D**). The advantage of both BA.4 and BA.5 over BA.2 was significantly lower at 27.9% and 48.5%, respectively. While BA.5 had a slight fitness advantage of 10% over BA.4 during the emergence of both lineages, this advantage was not sufficient to fully outcompete BA.4 for several months. However, late-stage BA.5 sub-lineages such as BQ.1.1 and BF.7 (**Table 1**) were able to push out BA.4 before XBB emerged by late November. XBB replaced these late-stage BA.5 sequences with a fitness advantage of 28.5%.

By characterizing the growth rates during the emergence windows for the major Omicron lineages, we have shown high variance in lineage replacement dynamics. The lack of a trend in our R_t_ ratio analysis suggests the fitness advantages of incoming lineages are likely being influenced by several non-uniform factors that vary for different emergence windows. We next sought to further dissect these observations through exploring possible underlying drivers of the incoming lineages’ fitness advantages.

### Average inter- and intra-lineage viral copy numbers vary over time and only partially explain lineage replacement patterns

To explain the specific mechanisms by which variants gain advantages over another, we evaluated the fitness advantage of each variant as a combination of intrinsic transmissibility and immune escape. For the purpose of this study, we used proxies derived from data collected through our genomic surveillance program for each of these factors.

As a proxy for intrinsic transmissibility, we measured virus copies in nasopharyngeal swab material using RT-qPCR. We used the same assay (Vogels et al. 2021) for each sample processed for sequencing which enables us to compare average cycle threshold (Ct) values per variant across a total of 11,111 samples (**Fig 3A**). We also converted the RT-qPCR values to genome equivalents (**Supplementary figure 3**).

**Figure 3.**
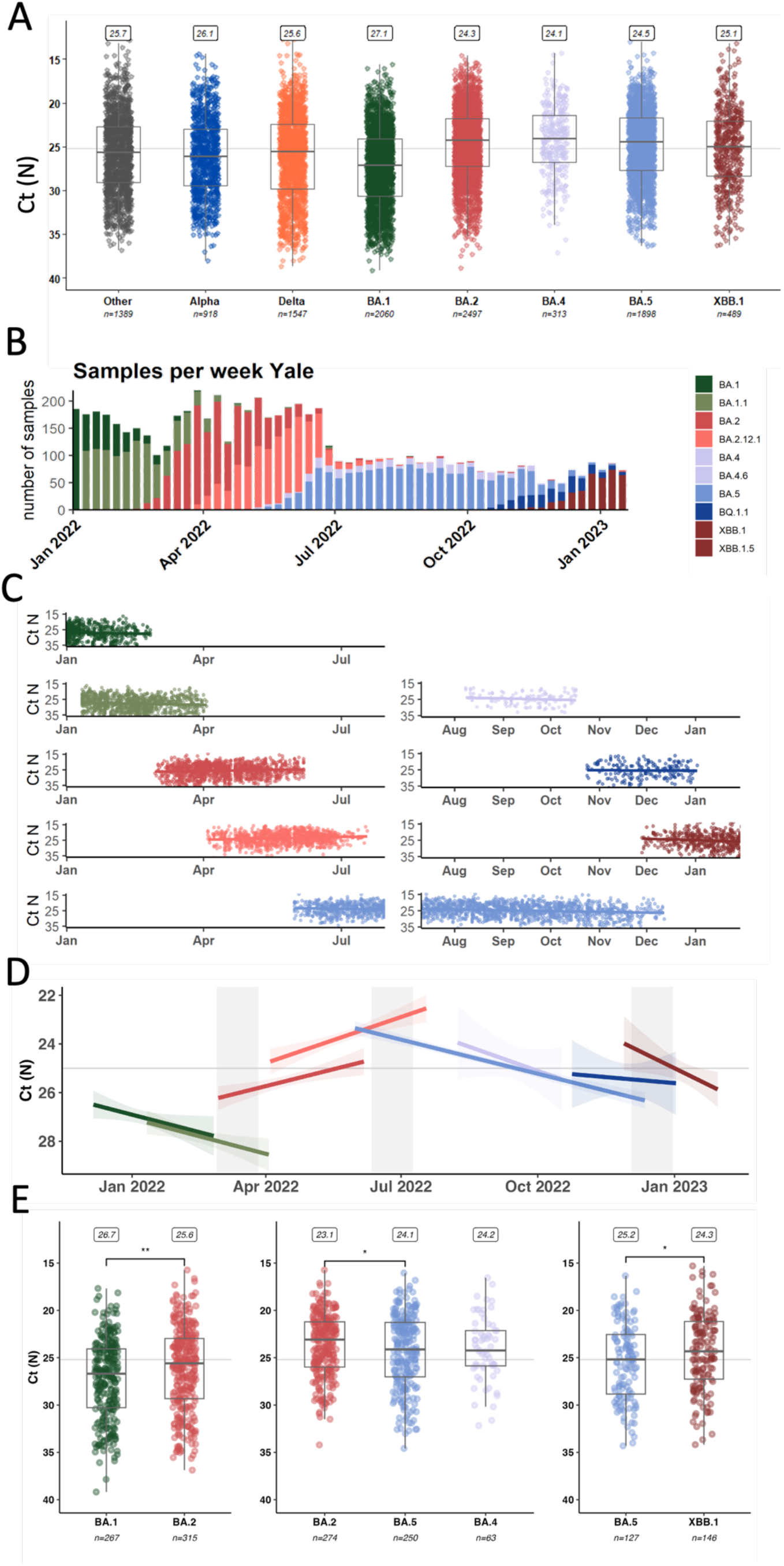
Comparing average qPCR Ct values as a proxy for variant intrinsic transmissibility. **(A)** Summary of all samples processed in our genomic surveillance programme from 2021 to January 2023 **(B)** Overview about the numbers and distribution of Omicron samples **(C)** Ct values from Omicron samples plotted together with modeled average over time **(D)** Based on (C), summary values for each of the Omicron lineages with emergence periods highlighted **(E)** Statistical analysis of Ct values retrieved from samples collected during the emergence periods.

Using cross-sectional data has caveats and is sensitive to several epidemiological and viral factors (Fryer et al. 2023; Hay et al. 2021). Changes in tissue tropism, symptom severity, and test-seeking behavior are expected to differ for Omicron infections compared to infections with pre-Omicron lineages. In the following, we are thus focusing on Omicron-lineage samples only.

For this, we analyzed 6,856 samples identified as Omicron-lineages (**Fig 3B**) collected from a wide range of individuals and disease statuses, including asymptomatic individuals detected through baseline surveillance of outpatients as well as inpatients and emergency department visits. We were particularly interested in Ct-value trends over time and plotted the raw data with temporal resolution (**Fig 3C)**. We then tested the data for the assumptions of linearity and fit linear regressions **(Fig 3D)** to derive coefficients as a measure of Ct-value change over time for each lineage (**Table 2**).

**Table 2.**
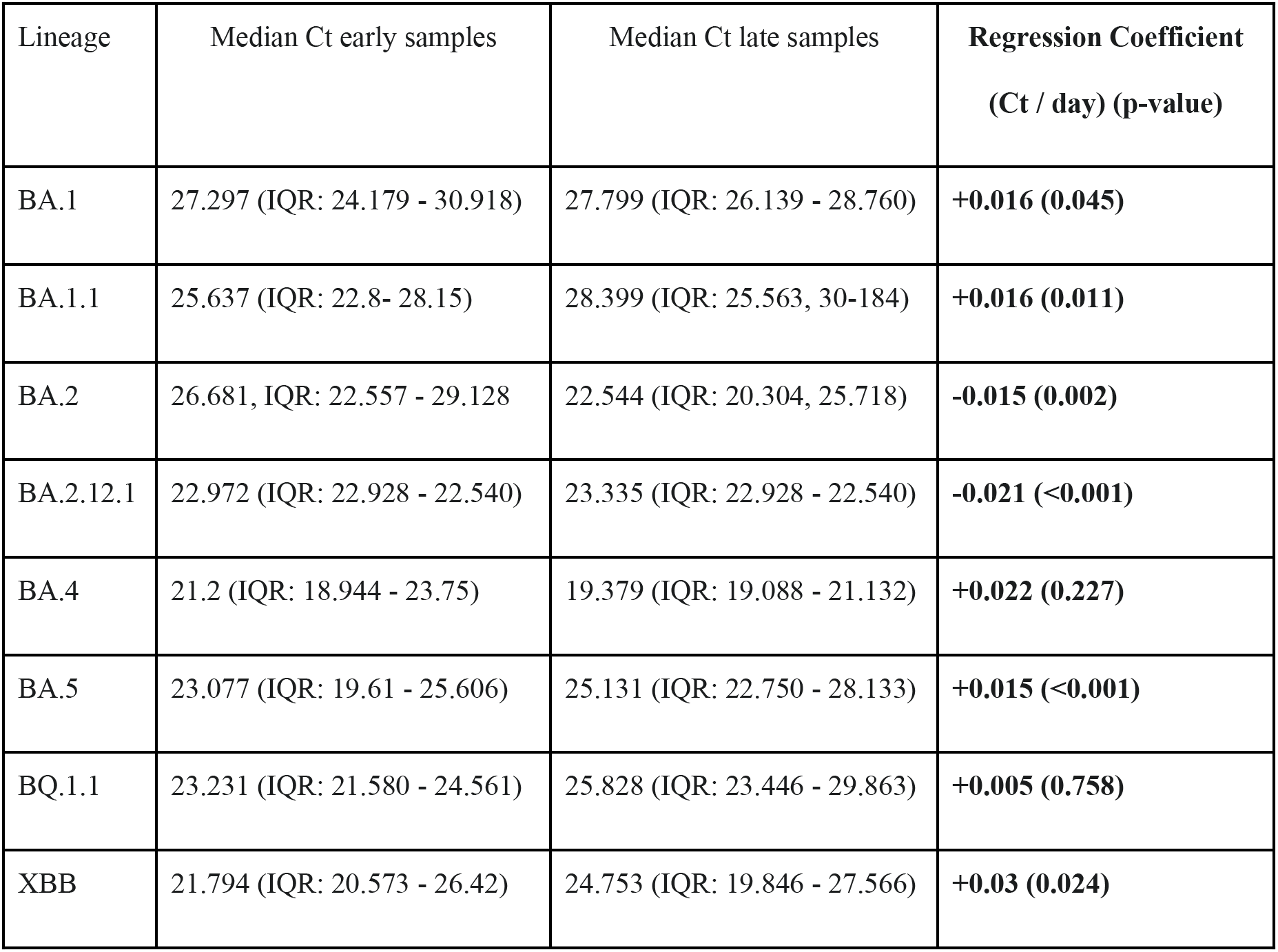
Trends in median Ct values of the major Omicron lineages over time.

Looking at trends over time, we noticed that the average Ct value for BA.1, BA.4, BA.5, and XBB samples increased over time (decreasing viral copy numbers) towards the end of each wave with a rate around 0.015-0.03 Ct values/day (**Table 2**). However, for the BA.2 and BA.2.12.1 samples, we observed a decreasing trend towards the end of the BA.2 wave with a decrease of 0.015 and 0.021 Ct values/day.

Next, we compared only samples that were collected specifically during the emergence period of an incoming lineage, defined as 14 days on either side of the date where the previously dominant and the incoming variant each account for 50% of the samples (**Supplement Table 2**).

Early BA.2 samples had significantly lower Ct values (median 25.6, Interquartile Range (IQR) 23.0-29.3) compared to late BA.1 samples (median 26.7, IQR 24.1-30.3) (p-value: 0.003) (**Fig 3E**). This supports previous reports from two European surveillance programs that also showed higher viral copies of BA.2 over BA.1 as measured by qPCR (Lentini et al. 2022; Musalkova et al. 2023). When looking at the emergence period of BA.4 and BA.5 compared to late-stage BA.2 samples, samples infected with the incoming variant had significantly higher Ct values than BA.2 samples (BA.4 median 24.2, IQR 22.1-25,9; BA.5 median 24.1, IQR 21.3-27.0, BA.2 median 23.1, IQR 21.2-26.0) from that same period (p-value: 0.034). There was no statistically significant difference in the Ct values between BA.4 and BA.5 samples (p-value: 0.829). Finally, early XBB samples again had lower Ct values (median 24.3, IQR 21.2-27.3) compared to late-stage BA.5 samples (including BQ.1.1) (median 25.2, IQR 22.5-28.8) (p-value: 0.033).

We also sought to investigate whether there was any association of patient metadata with Ct value in these first XBB cases by stratifying the samples according to age, sex, vaccination status, and patient class (**Supplementary Figure 4**). There was no significant difference in Ct values based on vaccination status. However, we found that people admitted to the emergency department had, on average, lower Ct values compared to those tested as inpatients or outpatients.

In summary, using an RT-qPCR assay for assessing viral copy number for each sample we submitted to whole-genome sequencing, we revealed striking temporal patterns. A higher viral copy number was not necessarily decisive for a lineage’s fitness advantage as, for example, incoming BA.5 samples had lower viral copy numbers than late BA.2 samples. Together, this hints at a varying advantage conferred by enhanced intrinsic transmissibility, as measured by viral copy numbers, that changed over the course of the year and seems mostly linked to BA.2-derived lineages.

### Likelihood of incoming lineages to cause breakthrough infections in recent vaccinees varies across different Omicron lineages

As BA.4 and BA.5 replaced BA.2 without clear advantage in their viral copy numbers, we next sought to assess the immune escape capacity to circumvent vaccine-induced immunity as another potential feature to increase a variant’s fitness.

A blood donor seroprevalence survey showed that in December 2021, 95.5% of donors (95% Confidence Interval (CI) 93.5-96.9%) had antibodies against the Spike antigen, compared to 17.8% (CI 15.3-20.5%) with infection-induced antibodies (Busch et al. 2022; Jones et al. 2022). The US CDC reports high vaccination coverage in Connecticut (70% coverage with 2 doses in September 2021) as well as uptake of a third dose booster towards the end of 2021 (55% in January 2022) (**Fig 4A**). Further, bi-valent booster shots (Wuhan-Hu-1+BA.5) were administered to 23% of the population (**Fig 4A**). During 2022, the estimated proportion of individuals with at least one previous SARS-CoV-2 infection passed 75% in July (Klaassen et al. 2022, 2023) (**Fig 4B**). Thus, the pool of antigen-naïve hosts was greatly depleted towards mid-2022. Consequently, the fitness advantage of variants being able to escape from this high population immunity were expected to increase over time (**Fig 4B**). Accordingly, in experimental assays with serum collected from curated cohorts with defined antigen exposures, Omicron lineages BA.1, BA.2, BA.5 and XBB showed an astounding sequential drop in sensitivity to neutralization by vaccine- and infection induced antibodies (Cao et al. 2023; Mykytyn et al. 2023a,b; Rössler et al. 2022).

**Figure 4.**
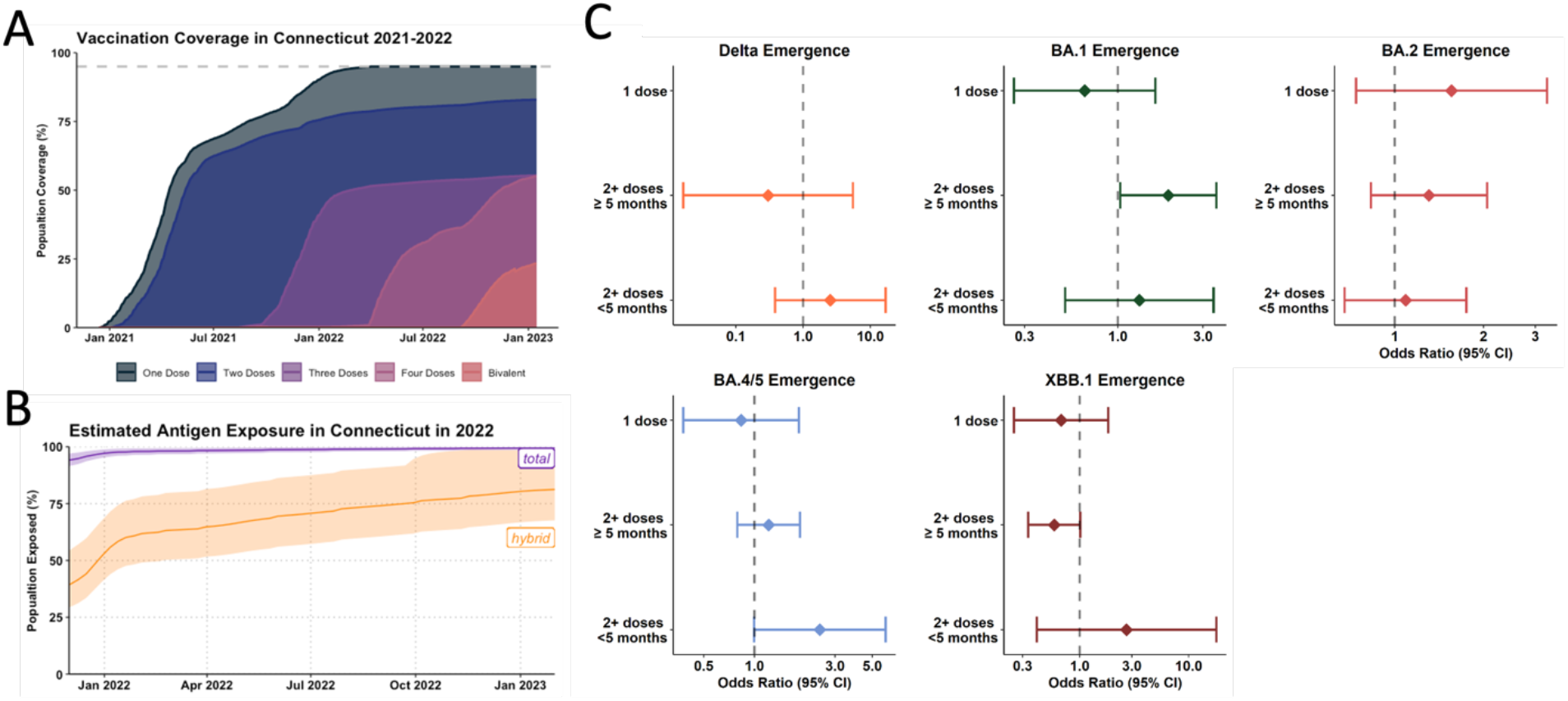
Influence of vaccine uptake and community-immunity levels on Omicron lineage emergence. **(A)** Vaccination coverage of Connecticut as reported by the US CDC for 2021 and 2022 **(B)** Estimated antigen exposure based on estimates from covidestim model according to either total (vaccination and/or infection) (purple) and hybrid exposure (infection and/or vaccination) (orange) **(C)** Logistic regression model to compare the ratios of infections caused by the incoming or previously dominant variant based on vaccination status during the emergence windows. BA.4 and BA.5 were analyzed together due to the similarity in the Spike region.

To examine how immune escape contributed to the fitness advantages of incoming Omicron lineages, we compared the likelihood of the incoming lineage to cause more vaccine breakthrough infections (BTI) than the dominant lineage during a five-week window of variant emergence (**Supplement Table 3**). We fitted mixed-effect multivariable logistic regression models adjusted for several potential confounding variables, including vaccination status, sex, age, location, and calendar time.

We show an increased odds of being infected with BA.1 compared to Delta amongst those vaccinated with at least 2 doses and who were more than 5 months from their most recent vaccination (OR: 1.92, 95% CI: 1.03 - 3.56) **(Fig 4C)**. Thus, enhanced immune escape played a role in the emergence of BA.1 while not being a significant driver of fitness advantage in the emergence of Delta in 2021. This result is in line with our previous study based on a similar population that found enhanced odds of being infected with Omicron BA.1 in triple-vaccinated individuals (Chaguza et al. 2022).

For the emergence of BA.2, we did not find overrepresentation in causing breakthrough infections during its emergence period, likely suggesting little to no effect of immune escape in our model **(Figure 4C)**. During the emergence of BA.5, we found a tendency for an increased risk of having been infected with BA.5 vs. BA.2 when having been vaccinated in the last 5 months prior to infection (OR: 2.441, 95% CI: 0.995 - 5.988), though this did not reach statistical significance **(Figure 4C)**. Finally, for the emergence of XBB, we found a tendency for a decreased likelihood of being infected with XBB amongst those who are more than 5 months from their most recent vaccination (OR: 0.586, 95% CI: 0.339-1.013) albeit this did not reach statistical significance in our analysis **(Figure 4C)**.

While these models are limited in understanding vaccine breakthrough infections from a population-level rather than mechanistic perspective, various in-vitro studies lend support to and provide context for our findings. Notably, the seeming lack of vaccine effects in the emergence of BA.2 can be explained by the low antigenic distance between BA.2 and BA.1, especially in people having received 3 monovalent doses (Arora et al. 2022b; Hoffmann et al. 2022; Kurhade et al. 2022; Tan et al. 2022). Additionally, several vaccine cohort studies have shown that BA.5 is more immune-evasive than BA.2 (Arora et al. 2022a; Cao et al. 2022b; Hoffmann et al. 2023b). The lack of statistical significance seen for the emergence of BA.5 in our analysis may be attributable to the comparatively small sample size for these strata in our dataset **(Supplement Table 10)**.

Our analysis shows that the advantage an emerging lineage gained through immune escape varies over time and that in-vitro findings may not fully translate to a heterologous immunity landscape influenced by individual combinations of infections, boosters, shifts in test-seeking behavior, and other population-level factors.

### A conceptual model describing Omicron lineage fitness in a highly dynamic fitness landscape

Lastly, we explored how the infection dynamics of each variant influenced the emergence of the subsequent replacement variant. For this, we analyzed population-level immune exposure by calculating the number of infections 60-, 90-, or 120-days prior to the time point where an incoming lineage reached 5% of sequence frequency (**Fig 5A**). Emerging BA.2 lineages in March 2022 faced a population that experienced around 1.5 million BA.1 infections within the last 90 days, the time frame where antibody-mediated protection against re-infections is expected to be highest. For the emergence of BA.5 and XBB, this figure was around 0.7 million. This extensive infection dynamic led to a gradual rise in the proportion of Connecticut’s population that was protected against reinfection as estimated through the covidestim model (**Fig 5B)** (Klaassen et al. 2022, 2023). Based on these estimates, we schematically plotted the relative importance of either intrinsic transmissibility or antigenic distance for an emerging variant’s fitness advantage over the course of 2022 (**Fig 5C).** While immune escape increases gradually, following the number of people with Omicron antigen exposure, the advantage conferred through higher intrinsic transmissibility mainly played a role after the BA.1 and BA.5 waves, which infected up to 50% of the population each.

**Figure 5.**
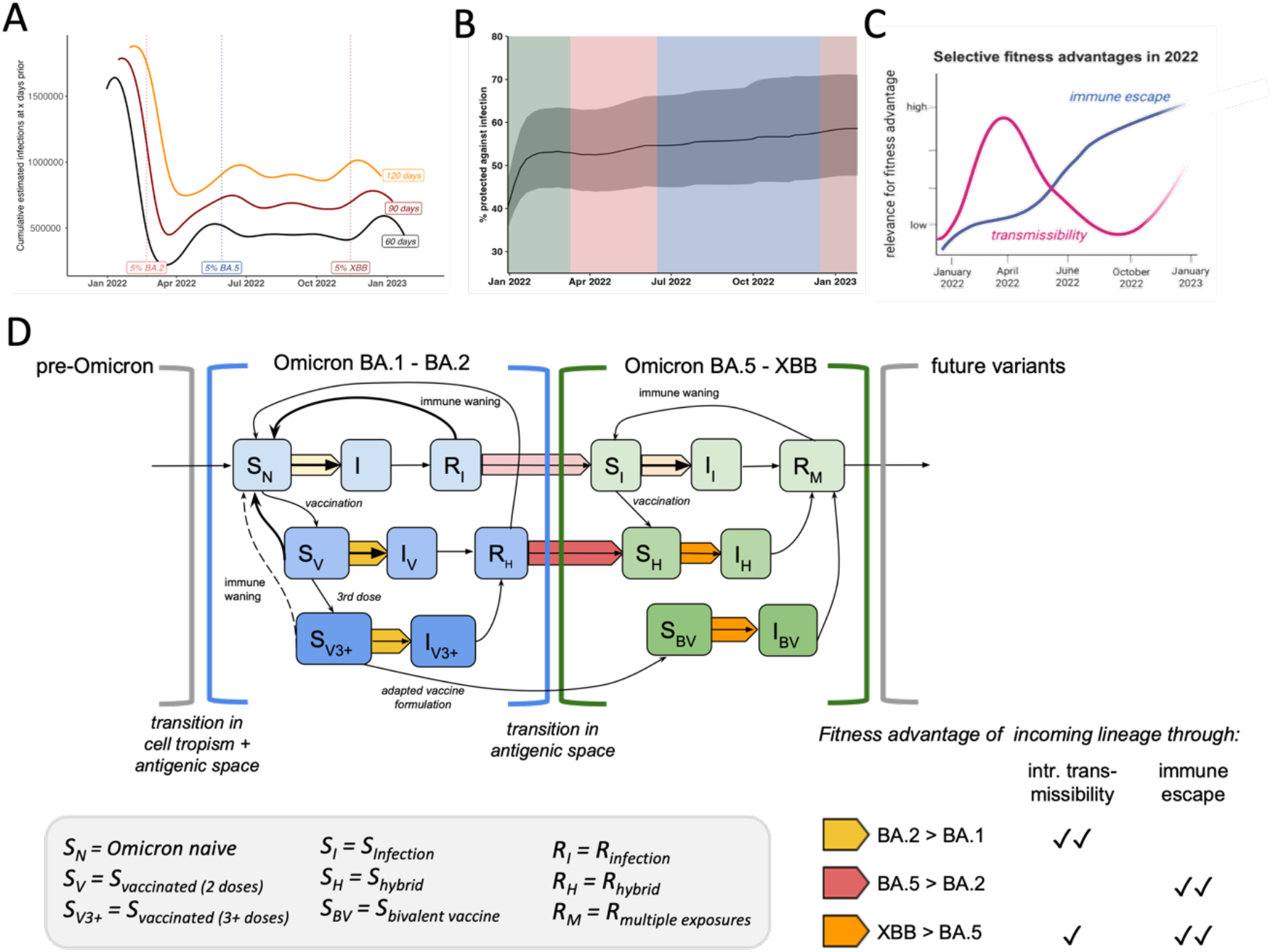
Summary of selective fitness advantages of Omicron lineage emergence in a dynamic host immunity landscape. **(A)** Sliding-window analysis of cumulative estimated infections 60, 90 or 120 days prior to each date on the plot with highlights when emerging Omicron lineages reached 5% of total samples **(B)** Frequency of estimated % of the population thought to be protected against infection based on vaccine and or infection status based on the covidestim model **(C)** Conceptual visualization of selective advantages of incoming Omicron lineages over the year **(D)** SIR-based conceptual transmission model highlighting different fitness advantages of incoming Omicron lineages depending on the status of the host population. Shades of yellow, orange and red represent the different combinations of advantageous fitness traits as outlined in the table in the right corner.

Summarizing our findings presented here, together with evidence from the literature, we developed a conceptual model based on a classical susceptible, infected, and recovered (SIR) compartmental dynamic transmission model highlighting fitness determinants of Omicron lineages (**Fig 5D**). Since Omicron BA.1 encompassed a vastly divergent Spike protein from pre-Omicron viruses, the previously observed high efficacy of vaccines, especially mRNA-platform based formulations, was greatly diminished (Andrews et al. 2022; Cao et al. 2022; Cele et al. 2022; Garcia-Beltran et al. 2022). In the model, this is reflected by opening a novel antigenic space (also defined as serotype) compared to previous pre-Omicron lineages (Simon-Loriere & Schwartz 2022). We include three different levels of susceptibility depending on vaccine status (S_N_, S_V_, S_V3+_), as these strata have shown to harbor different quantities and qualities in antibody levels and thus protection against a first Omicron infection (Muecksch et al. 2022; Park et al. 2022; Schaefer-Babajew et al. 2022). The exploration of this new antigenic space confers a fitness advantage in itself, moving 40-50% of Connecticut’s population from the ‘susceptible’ (S_N_ = Omicron-naive, i.e. no previous Omicron antigen exposure) compartment to ‘recovered’ (R_I_ for unvaccinated or R_H_ for vaccinated).

The fraction of the population that just recovered from BA.1 (**Fig 5A**) was likely fairly well protected against a swift reinfection with either BA.1 or the antigenically similar BA.2 (Zhou et al. 2023). Based on epidemiological data, the transition from the R_I_/R_H_ bin back to the ‘susceptible’ bin within the same antigenic space takes around 6 months (Andeweg et al. 2023; Malato et al. 2023; 2023). BA.2 was competing with BA.1 to infect the remaining ‘susceptible’ population (i.e. Omicron-naive S_N_) fraction of the population. Based on our analysis, we conclude BA.2 outcompeted BA.1 in reaching this susceptible population faster through its higher intrinsic transmissibility (yellow arrow) (**Figs 3E and 5D**).

After the BA.1 and BA.2 waves, approximately one-third of the population remained in the Omicron-naive compartment (**Fig 1D**). The fitness advantage of variants in the same antigenic space, even with higher transmissibility (like BA.2.12.1), was restricted to this remaining pool of susceptibles. BA.4 and BA.5 successfully escaped from immunity conferred through pre-Omicron antigen contacts as well as BA.1/BA.2 infections and thus opened a second antigenic space (**Fig 5D**). Vaccinated or previously infected individuals fall back from being fairly protected (‘recovered’ bin) in the previous antigenic space to the ‘susceptible’ (S_I_, S_H_) state in the second antigenic space (**Fig 5D**). Although BA.4 and BA.5 displayed lower intrinsic transmissibility compared to BA.2 (**Fig 3D**), these variants outcompeted BA.2 as they could spread in a larger fraction of people (the R_I_ & R_H_ bins) (red arrow) (**Fig 5D**). The fairly high and steady transmission levels of BA.4 and 5 (**Fig 1C**) were likely defined by the rate of individuals moving from the recovered bin of the first antigen space to the S compartment in the second antigenic space (**Fig 5D**).

At the time XBB emerged, over 80% of the population had at least one Omicron antigen exposure (**Fig 4B**). For the purpose of our model, we assume that XBB covers a similar antigenic niche as BA.5. Although XBB displays immune evasion from BA.5-induced immunity, some cross-neutralization, especially in individuals with previous Omicron antigen contact, was reported (Kurhade et al. 2023; Springer et al. 2023). In order to outcompete the ongoing BA.5 transmission chains (**Fig 2D**), any incoming lineage had to maintain at least similar or higher levels of immune escape while also being more transmissible (orange arrow) (**Fig 5D**). Thus, XBB outcompeted late-stage BA.5 samples through its extensive immune escape profile (Hoffmann et al. 2023) paired with higher intrinsic transmissibility (**Fig 3E**) and thus spreading faster within the susceptibles in the second antigenic space (**Fig 5D**).

In summary, we provide a framework to assess critical parameters that define variant fitness of Omicron lineages in 2022. Ongoing SARS-CoV-2 evolution led to the emergence of lineages with greater flexibility in exploring different fitness niches in a highly dynamic host population through a differential combination of intrinsic transmissibility and immune escape.

## Discussion

We evaluated the underlying drivers of fitness advantages that led to the sequential sweeps of Omicron lineages in Connecticut during 2022. By determining selective traits that best explain the observed lineage dynamics, our analysis revealed the ability of SARS-CoV-2 lineages to thrive in a variety of highly dynamic fitness landscapes. A key result from the data presented here is that SARS-CoV-2 fitness was not dictated by a single factor, but rather a fine-tuned combination of intrinsic transmissibility and immune escape. This intrinsic volatility of the fitness landscape is, by definition, coupled with the antigen exposure history of the host population, making fitness advantages moving targets.

While novel virus lineages are designated according to their phylogenetic identity, inferring epidemiological relevance and antigenic space from genetic data alone remains challenging. Thus, in addition to continuing efforts of genomic surveillance, it remains crucial to collect relevant clinical and epidemiological patient data, ideally coupled with broad-scale and representative sero-prevalence surveys. Being able to link genetic information with additional laboratory and epidemiological data of locally matched host-pathogen populations and their epidemic outcomes is crucial for evaluating the potential of novel variants (Meijers et al. 2023).

2022 was unique in terms of the spread of the Omicron family, followed by highly synchronized antigen exposures resulting in fairly homogenous population-level immunity. Going forward, the immunity landscape will likely greatly diversify depending on individuals’ antigen exposure histories and on the synchronicity of waning rates between individuals. This has implications for future SARS-CoV-2 transmission as a significant portion of the population might become susceptible to newly circulating lineages entering the northern hemisphere winter season of 2023/2024. We showed that in 2022, high levels of population immunity towards the end of the year slowed down the growth rates of BA.5 and XBB. Thus, variant-turnovers will take longer in future and might be more subtle than the rapid lineage displacements in 2022. Additionally, the high number of co-circulating XBB-variants might further limit the growth rates of newly emerging variants (Beesley et al. 2023).

We demonstrate the utility of an integrated approach to genomic surveillance data collection for identifying key characteristics of SARS-CoV-2 fitness dynamics. Data linkage to key patient metadata was crucial to evaluate existing data from laboratory studies and curated vaccinee cohorts in a real-world, population-level setting and should be considered when designing surveillance systems. Such frameworks provide critical infrastructure to evaluate emerging lineages of known and unknown pathogens and can be further incorporated into tailoring public health responses and informing vaccine formulations. Lessons learned from SARS-CoV-2 can be utilized and further adapted to other pathogens to maximize the usefulness of such surveillance programmes for community health.

This retrospective analysis re-visits the lineage replacement dynamics of Omicron lineages that dominated Connecticut in 2022 and sheds light on the underlying drivers of their sequential fitness advantages. This study exemplifies a use-case for combining sequencing data with epidemiological and laboratory metadata to better understand pathogen emergence. Based on the high-level data used in this study, we showcase that linking vaccination status and viral copy numbers to genomic surveillance data may provide crucial tools to evaluate variant fitness for SARS-CoV-2 and beyond.

## Limitations

One limitation of this study might be the validity of these findings to other populations and settings. As outlined above, variant transmission advantages are intrinsically coupled to the local host population and thus our results are most applicable to other places with similar population characteristics, vaccination uptake rates, and public health policies. Whereas the specific dynamics of the different Omicron waves may vary locally, the data presented here is valuable for examining overall trends that are broadly applicable to other settings. Overall fitness advantages of each new Omicron lineage over their respective predecessor were fairly consistent globally as recently shown when analyzing different settings globally (Meijers et al. 2023).

A further potential limitation is that our dataset for Ct value comparison is based on a pre-selection from the overall pool of available SARS-CoV-2 positive samples based on a Ct value cut-off in the diagnostic test that were expected to yield high sequencing coverage. We think this reflects the proportion of samples that would possess the viral burden that is relevant for onward transmission and thus suitable for the purpose of this study. Further, it could be argued that Ct values do not necessarily scale with the amounts of infectious viral particles and that the kinetics of RNA clearance may not be directly related to clearance of infectious virus. For our specific assay, we used a standard curve to convert Ct values to absolute viral genome copies and we are usually able to isolate infectious virus from samples with Ct values below 30 (based on internal data). Further, symptom severity is thought to have remained similar over all Omicron lineages, reducing the potential for bias due to differences in test seeking behavior. Lastly, for the purposes of this study, we were interested in comparing trends between lineages in contrast to absolute values.

## Materials & Methods

### Ethics statement

The Institutional Review Board from the Yale University Human Research Protection Program determined that the RT-qPCR testing and sequencing of de-identified remnant COVID-19 clinical samples obtained from clinical partners conducted in this study is not research involving human subjects (IRB Protocol ID: 2000028599).

## Data sources

### Clinical sample collection, RT-qPCR, and sequencing

SARS-CoV-2 positive samples (nasal swabs in viral transport media) were collected through the Yale New Haven Hospital (YNHH) System as a part of routine inpatient and outpatient testing and sent to the Yale SARS-CoV-2 Genomic Surveillance Initiative. Using the MagMAX viral/pathogen nucleic acid isolation kit, nucleic acid was extracted from 300μl of each clinical sample and eluted into 75μl of elution buffer. Extracted nucleic acid was then tested using a “research use only” (RUO) RT-qPCR assay (Vogels et al. 2021) for SARS-CoV-2 RNA. Libraries were prepared for sequencing using the Illumina COVIDSeq Test (RUO version) and quantified using the Qubit High Sensitivity dsDNA kit. Negative controls were included for RNA extraction, cDNA synthesis, and amplicon generation.

Prepared libraries were sequenced at the Yale Center for Genomic Analysis on the Illumina NovaSeq with a 2x150 approach and at least 1 million reads per sample.

Reads were then aligned to the Wuhan-Hu-1 reference genome (GenBank MN908937.3) using BWA-MEM v.0.7.15 (Li 2013). Adaptor sequences were then trimmed, primer sequences masked, and consensus genomes called (simple majority >60% frequency) using iVar v1.3.133 (Grubaugh et al. 2019) and SAMtools(Danecek et al. 2021). When <20 reads were present at a site an ambiguous “N” was used, with negative controls consisting of ≥99% Ns. The Pangolin lineage assignment tool (O’Toole et al. 2021) was used for assigning viral lineages.

### Clinical metadata

We obtained patient metadata and vaccination records from the YNHH system and the Center for Outcomes Research and Evaluation (CORE) and matched these records to sequencing data through unique sample identifiers. Duplicate patient records or those with missing or inconsistent vaccination data were removed. We determined vaccination status at time of infection by comparing the sample collection date to the patient’s vaccination record dates.

We then categorized vaccine breakthrough statuses with respect to both the number of vaccine doses received more than 14 days prior to the collection date and the timing of the most recent vaccination relative to a 5 month period of time. Patient vaccination statuses at time of infection were thus categorized as: non-vaccine breakthrough, one dose vaccine breakthrough, two or more dose vaccine breakthrough greater than 5 months since the most recent vaccination, or two or more dose vaccine breakthrough within 5 months since the most recent vaccination.

### Population vaccination trends

We obtained data on vaccination trends in Connecticut from the Centers for Disease Control and Prevention (CDC) (CDC 2021).

### Population variant trends

We obtained variant trend data for Connecticut from the Global Initiative on Sharing All Influenza Data (GISAID).

### Variant Rt and immunity estimates

We obtained variant cumulative estimated case counts from covidestim, a Bayesian nowcasting approach that incorporates reported cases, hospitalizations, immunity (Klaassen et al. 2022, 2023), exposures, and vaccination data to generate state and county level estimates of variant specific infections (Chitwood et al. 2022). To obtain variant-specific Rt estimates, we used the variant-specific infection estimates from covidestim with the EpiEstim R package (Nouvellet et al. 2018).

## Analyses

### Variant Rt ratios

To compare R_t_ values between variants, we first selected 14-day periods when a new variant was emerging in the population. For each of these time periods, we then divided the daily R_t_ value of the emerging variant by the R_t_ value of the established variant for the same calendar day to calculate daily R_t_ ratios.

### Variant emergence periods and logistic growth rates

To determine the length of time each variant needed to become established in the population, we calculated daily frequencies for each variant across Connecticut using case count data from GISAID. We then defined a variant’s emergence period as the date from when a variant first accounted for 5% of all cases to the first date the variant reached its maximum frequency in the population. We then fitted a locally estimated scatterplot smoothing (loess) curve to the data and extracted the fitted value corresponding to a frequency of 50% in the population. With this value we determined the number of days it took each variant to increase from a frequency of 5% in the population to 50%.

Using the same emergence periods, we fitted binomial logistic regression models where the variant lineage was modeled as a function of calendar time. For each model, the variant(s) being displaced in the population served as the reference group. We then extracted the coefficient of the predictor variable of each model to determine the logistic growth rate for each variant.

### Variant Ct values over time and in periods of emergence

To understand how Ct values change across time, we subset each variant to the period of time where it was above 10% frequency in the population using RT-qPCR data from the Yale SARS-CoV-2 Genomic Surveillance Initiative. For each of these time periods and variants we tested for heteroscedasticity via a Breusch-Pagan test, checked for non-linearity or outlier values by plotting the residual values against the fitted values, and tested for normality via a Q-Q plot and frequency histogram of model residuals. We then fitted linear regression models where the Ct value was modeled as a function of calendar time.

To compare Ct values between variants, we identified 3, 4-week periods of variant co-circulation using the same dataset. For the two periods of pairwise comparisons, we performed Wilcoxon rank sum tests. For the comparison between three variants, we performed a Welch’s ANOVA test as well as a one-way ANOVA test to test for concordance, followed by post-hoc pairwise tests via Tukey’s honest significance test.

### Mixed effect multivariable logistic regression models

To determine the impact of vaccinations in periods of variant emergence, we used sequencing data from the Yale SARS-CoV-2 Genomic Surveillance Initiative matched to vaccination data from the YNHH System and CORE to identify five, five-week periods of variant co-circulation. We selected the specific date ranges of these periods so as to balance the number of unvaccinated individuals attributed to each variant in each period.

For these five periods we then fit mixed effect multivariable logistic regression models with a dichotomous outcome of the co-circulating variants found in each time period. To dichotomize the outcome in periods when more than two variants were circulating, we aggregated variants that emerged or were displaced contemporaneously, with the reference set as the variant(s) that was being displaced in the population.

Model covariates were selected via an Akaike information criterion (AIC) selection criteria test and included vaccination status at the time of infection, patient sex (male or female), age (5-17, 18-39, 40-64, 65+), town of residence as a random effect, and calendar time as a linear predictor. Non-vaccine breakthrough, male sex, and the 18-39 age group served as variable reference levels. Due to the inability for under 5 years olds to receive vaccinations for the majority of our study period, we restricted our analysis to individuals 5 years and older.

To test the impact of the date interval lengths, we performed a sensitivity analysis by modifying the period of emergence from 5 weeks to 3,4,6,7, and 8 weeks and found minimal differences in the results.

### Factors impacting XBB.1.5 Ct values

To investigate the factors that impact Ct values associated with the XBB.1 variant infections, we subset sequencing data from the Yale SARS-CoV-2 Genomic Surveillance Initiative to only those infections caused by lineages within the XBB.1 parent lineage. We then identified calendar time, sex (female or male), patient class (inpatient, outpatient, or emergency), age (<18, 18-49, 50-69, 70+), and vaccination status at time of infection as variables that could impact Ct values.

To understand the impact of calendar time, we tested for heteroscedasticity via a Breusch-Pagan test, checked for non-linearity or outlier values by plotting the residual values against the fitted values, and tested for normality via a Q-Q plot and frequency histogram of model residuals. We then fit a linear regression model to the data with Ct value as a function of calendar time.

For the remaining variables we assessed the normality of the Ct value distributions via Shapiro-Wilk tests and the variance of the Ct value distributions via Bartlett or F-tests. To test for differences in Ct values by sex, we performed equal variance t-tests. For the remaining variables, we performed Welch’s ANOVA tests as well as one-way ANOVA tests to test for concordance, followed by post-hoc pairwise comparisons via Tukey’s honest significance test.

### Conceptual SIR Model

To synthesize our findings, we constructed a conceptual framework of variant fitness based on the traditional SIR transmission model. As a conceptual framework, this model does not attempt to explicitly simulate transmission through quantitative approaches. Rather, the model provides a visual representation of the proposed mechanisms by which the Omicron lineages may have gained selective advantages based on the results of our other analyses and findings from the literature.

### Statistical analysis and data availability

We used the R statistical software (v. 4.2.1) (R Core Team (2021) R: A Language and Environment for Statistical Computing. R Foundation for Statistical Computing, Vienna.) for all statistical analysis and figures.

Data and code used in this study are publicly available on GitHub (https://github.com/NickChen10/Omicron_project).

## Data Availability

All data produced in the present study are available upon reasonable request to the authors.

## Acknowledgments

We gratefully acknowledge the work of all members of the Yale Genomic Surveillance Initiative especially Center for Outcome Research at Yale New Haven Hospital. We thank all authors from the originating laboratories responsible for obtaining the specimens, as well as the submitting laboratories where the genomic data were generated and shared via GISAID. This project is supported by the CDC Broad Agency Announcement Contracts 75D30122C14697 (awarded to NDG) and 75D30121C10273 (supports KF), the Connecticut Department of Public Health (CDPH) contract 21PSX0049 (awarded to the Yale Center for Genome Analysis and supports NDG), and the Council of State and Territorial Epidemiologists contract NU38OT000297 (supports KF). This work does not necessarily represent the views of the CDC or CDPH.

## Author contribution

Conceptualization: NC, NDG, AMH

Methodology: NC, KP, NDG, AMH

Investigation: NC, KP, AMH, DMW, JLW, VP, NDG, RL, FK

Visualization: NC, KP, AMH

Funding acquisition: NDG

Supervision: NDG, AMH

Writing – original draft: NC, AMH

Writing – review & editing: all authors

## Competing Interests

NDG is a paid consultant for BioNTech, DMW has received consulting fees from Pfizer, Merck, and GSK, unrelated to this manuscript, and has been PI on research grants from Pfizer and Merck to Yale, unrelated to this manuscript. JLW has received consulting fees from Pfizer and Revelar Biotherapeutics Inc unrelated to this manuscript.

## Notes

### Funding Statement

This project is supported by the CDC Broad Agency Announcement Contracts 75D30122C14697 and 75D30121C10273, the Connecticut Department of Public Health (CDPH) contract 21PSX0049, and the Council of State and Territorial Epidemiologists contract NU38OT000297. This work does not necessarily represent the views of the CDC or CDPH.

