## Supplementary Material for "Escalating combinations of enhanced infectivity and immune escape define SARS-CoV-2 Omicron lineage replacement"

### Supplement

**Supplementary Table 1 Emergence periods for Growth advantage calculation**

| Dominant Variant | Incoming Variant | Start | End |
|---|---|---|---|
| Delta | BA.1 | Dec 15 2021 | Dec 22 2021 |
| BA.1 | BA.2 | March 6 2022 | March 20 2022 |
| BA.2 | BA.4/5 | June 18 2022 | July 2 2022 |
| BA.5 | XBB | December 16 2022 | December 30 2022 |

**Supplementary Table 2 Emergence periods for Ct value comparison**

| Dominant Variant | Incoming Variant | Start | Crossover | End |
|---|---|---|---|---|
| BA.1 | BA.2 | February 27 2022 | March 13 2022 | March 27 2022 |
| BA.2 | BA.4/5 | June 11 2022 | June 25 2022 | July 09 2022 |
| BA.5 | XBB | December 03 2022 | December 17 2022 | December 31 2022 |

**Supplementary Table 3 Emergence periods for vaccine breakthrough infections comparison**

| Dominant Variant | Incoming Variant | Start | End |
|---|---|---|---|
| Delta | BA.1 | November 30 2021 | January 04 2022 |
| BA.1 | BA.2 | February 23 2022 | March 30 2022 |
| BA.2 | BA.4/5 | June 09 2022 | July 14 2022 |
| BA.5 | XBB | December 07 2022 | January 11 2023 |

#### Supplementary Table 4: Patient demographics by Vaccination Status (n = 14246)

| | Unvaccinated (n=7234) | 1 dose (n=972) | 2+ doses ≥ 5 months (n=4688) | 2+ doses < 5 months (n=1352) | Overall (n=14246) |
|---|---|---|---|---|---|
| **Age group** | | | | | |
| 5-17 | 2051 (28.4%) | 49 (5.0%) | 222 (4.7%) | 124 (9.2%) | 2446 (17.2%) |
| 18-39 | 2635 (36.4%) | 327 (33.6%) | 1197 (25.5%) | 403 (29.8%) | 4562 (32.0%) |
| 40-64 | 1999 (27.6%) | 453 (46.6%) | 2045 (43.6%) | 558 (41.3%) | 5055 (35.5%) |
| 65+ | 549 (7.6%) | 143 (14.7%) | 1224 (26.1%) | 267 (19.7%) | 2183 (15.3%) |
| **Sex** | | | | | |
| Female | 3273 (45.2%) | 464 (47.7%) | 2346 (50.0%) | 743 (55.0%) | 6826 (47.9%) |
| Male | 3157 (43.6%) | 413 (42.5%) | 1744 (37.2%) | 538 (39.8%) | 5852 (41.1%) |
| Other | 21 (0.3%) | 1 (0.1%) | 12 (0.3%) | 0 (0%) | 34 (0.2%) |
| Unknown | 783 (10.8%) | 94 (9.7%) | 586 (12.5%) | 71 (5.3%) | 1534 (10.8%) |

**Supplementary Table 5: Delta Emergence Model**

| Covariates | Odds ratio ($e^{\beta}$, 95% CI) | p-value |
|---|---|---|
| **Vaccination status** | | |
| Unvaccinated | Ref. | Ref. |
| One dose | NA | NA |
| 2+ doses ≥ 5 months | 0.302 (0.017, 5.481) | 0.418 |
| 2+ doses< 5 months | 2.533 (0.381, 16.818) | 0.336 |
| **Age category** | | |
| 18-39 | Ref. | Ref. |
| 5-17 | 0.573 (0.135, 2.444) | 0.452 |
| 40-64 | 1.106 (0.296, 4.131) | 0.881 |
| 65+ | 0.083 (0.008, 0.814) | 0.033 |
| **Sex** | | |
| Male | Ref. | Ref. |
| Female | 1.689 (0.562, 5.074) | 0.350 |
| **Time** | 3.898 (2.023, 7.511) | <0.001 |

**Supplementary Table 6: BA.1 Emergence Model**

| Covariates | Odds ratio ($e^{\beta}$, 95% CI) | p-value |
|---|---|---|
| **Vaccination status** | | |
| Unvaccinated | Ref. | Ref. |
| One dose | 0.651 (0.262, 1.621) | 0.357 |
| 2+ doses ≥ 5 months | 1.917 (1.033, 3.558) | 0.0393 |
| 2+ doses< 5 months | 1.320 (0.507, 3.440) | 0.570 |
| **Age category** | | |
| 18-39 | Ref. | Ref. |
| 5-17 | 0.280 (0.131, 0.598) | 0.001 |
| 40-64 | 0.471 (0.258, 0.861) | 0.014 |
| 65+ | 0.365 (0.146, 0.912) | 0.031 |
| **Sex** | | |
| Male | Ref. | Ref. |
| Female | 0.956 (0.574, 1.592) | 0.863 |
| **Time** | 50.430 (27.761, 91.611) | <0.001 |

**Supplementary Table 7: BA.2 Emergence Model**

| Covariates | Odds ratio ($e^{\beta}$, 95% CI) | p-value |
|---|---|---|
| **Vaccination status** | | |
| Unvaccinated | Ref. | Ref. |
| One dose | 1.558 (0.739, 3.285) | 0.244 |
| 2+ doses ≥ 5 months | 1.308 (0.830, 2.061) | 0.247 |
| 2+ doses< 5 months | 1.089 (0.677, 1.752) | 0.724 |
| **Age category** | | |
| 18-39 | Ref. | Ref. |
| 5-17 | 1.087 (0.614, 1.926) | 0.775 |
| 40-64 | 1.003 (0.652, 1.542) | 0.989 |
| 65+ | 1.251 (0.673, 2.327) | 0.479 |
| **Sex** | | |
| Male | Ref. | Ref. |
| Female | 1.066 (0.745, 1.526) | 0.727 |
| **Time** | | <0.001 |

**Supplementary Table 8: BA.4/5 Emergence Model**

| Covariates | Odds ratio ($e^{\beta}$, 95% CI) | p-value |
|---|---|---|
| **Vaccination status** | | |
| Unvaccinated | Ref. | Ref. |
| One dose | 0.834 (0.380, 1.835) | 0.653 |
| 2+ doses ≥ 5 months | 1.214 (0.791, 1.863) | 0.374 |
| 2+ doses< 5 months | 2.441 (0.995, 5.988) | 0.051 |
| **Age category** | | |
| 18-39 | Ref. | Ref. |
| 5-17 | 0.782 (0.396, 1.546) | 0.480 |
| 40-64 | 0.858 (0.551, 1.335) | 0.497 |
| 65+ | 0.630 (0.373, 1.062) | 0.083 |
| **Time** | 2.924 (2.363, 3.617) | <0.001 |

**Supplementary Table 9: XBB.1.5 Emergence Model**

| Covariates | Odds ratio ($e^{\beta}$, 95% CI) | p-value |
|---|---|---|
| **Vaccination status** | | |
| Unvaccinated | Ref. | Ref. |
| One dose | 0.677 (0.251, 1.830) | 0.442 |
| 2+ doses ≥ 5 months | 0.586 (0.339, 1.013) | 0.056 |
| 2+ doses< 5 months | 2.690 (0.405, 17.859) | 0.306 |
| **Age category** | | |
| 18-39 | Ref. | Ref. |
| 5-17 | 0.538 (0.218, 1.326) | 0.178 |
| 40-64 | 0.575 (0.313, 1.056) | 0.075 |
| 65+ | 0.539 (0.290, 0.999) | 0.0497 |
| **Sex** | | |
| Male | Ref. | Ref. |
| Female | 1.372 (0.865, 2.177) | 0.179 |
| **Time** | 2.251 (1.765, 2.872) | <0.001 |

**Supplementary Table 10: Vaccination Strata by Emergence Period**

| Emerging Variant | **Delta** | **BA.1** | **BA.2** | **BA.4/5** | **XBB** |
|---|---|---|---|---|---|
| **Vaccination Status** | | | | | |
| Unvaccinated | 72 | 293 | 213 | 174 | 101 |
| One dose | 3 | 82 | 50 | 41 | 24 |
| 2+ doses ≥ 5 months | 2 | 376 | 234 | 378 | 240 |
| 2+ doses< 5 months | 9 | 78 | 181 | 36 | 6 |
| Total | 86 | 829 | 678 | 629 | 371 |

### Supplementary Figure 1

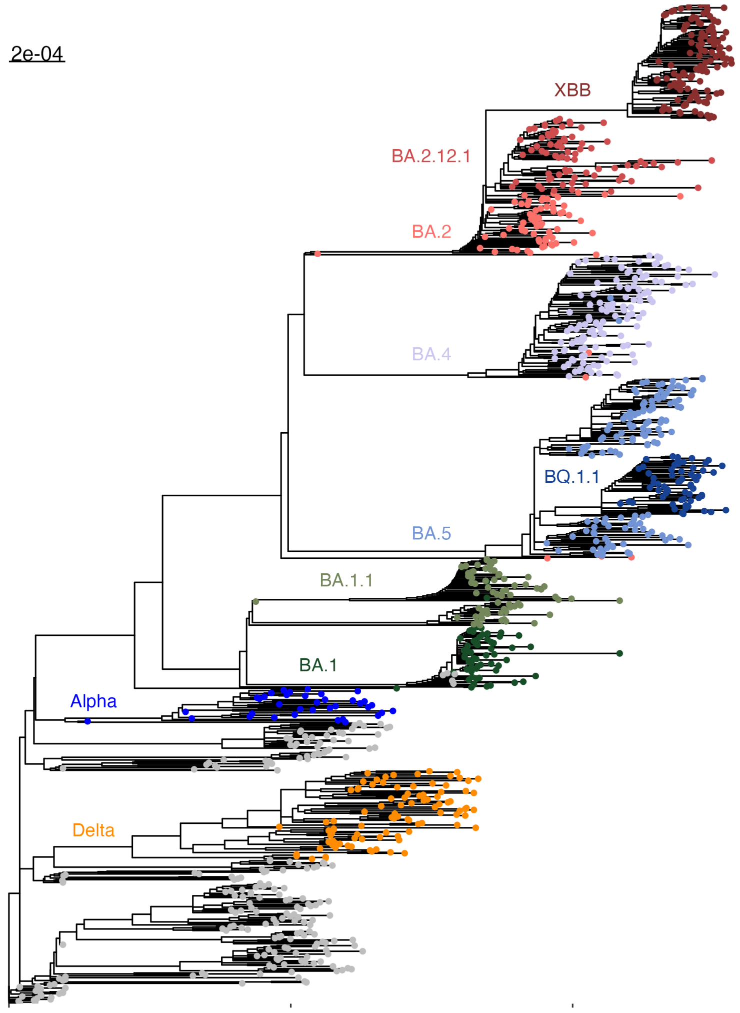


**Figure S1 Phylogenetic tree representing the genetic distance of different variants of concern as well as the Omicron lineages BA.1, BA.2, BA.4., BA.5 and XBB described in this study.** The tree is based on a sub-sampling of 1000 samples primarily subsampled from the Yale-SARS-CoV-2 genomic surveillance initiative including some international reference sequences. The tree is rooted in the Wuhan-Hu1 sample. Across the Omicron family, a remarkable degree of convergent intra-lineage evolution has led to the emergence of at least one major sub-lineage. This is most clearly exemplified in the independent acquisition of amino acid changes at the same positions in the Spike region (e.g. BA.1.1 (R346K), BA.4.6 (R346T) and BQ.1.1 (R346T)).

### Supplementary Figure 2

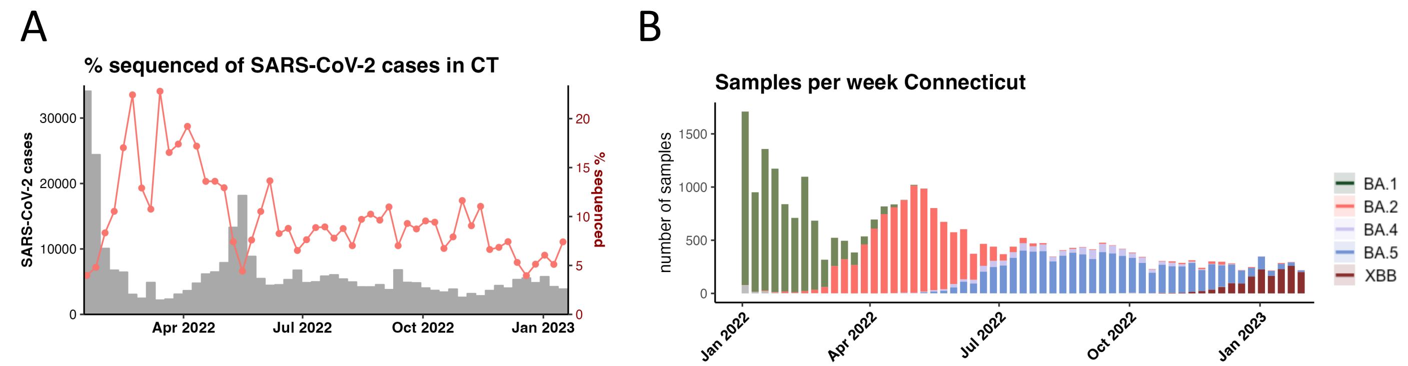


**Figure S2 Yale SARS-CoV-2 Genomic Surveillance Initiative in 2022 (A)** Frequency of % sequenced (red line) versus reported cases (grey bars) **(B)** Number and distribution of SARS-CoV-2 samples deposited on GISAID throughout 2022

### Supplementary Figure 3

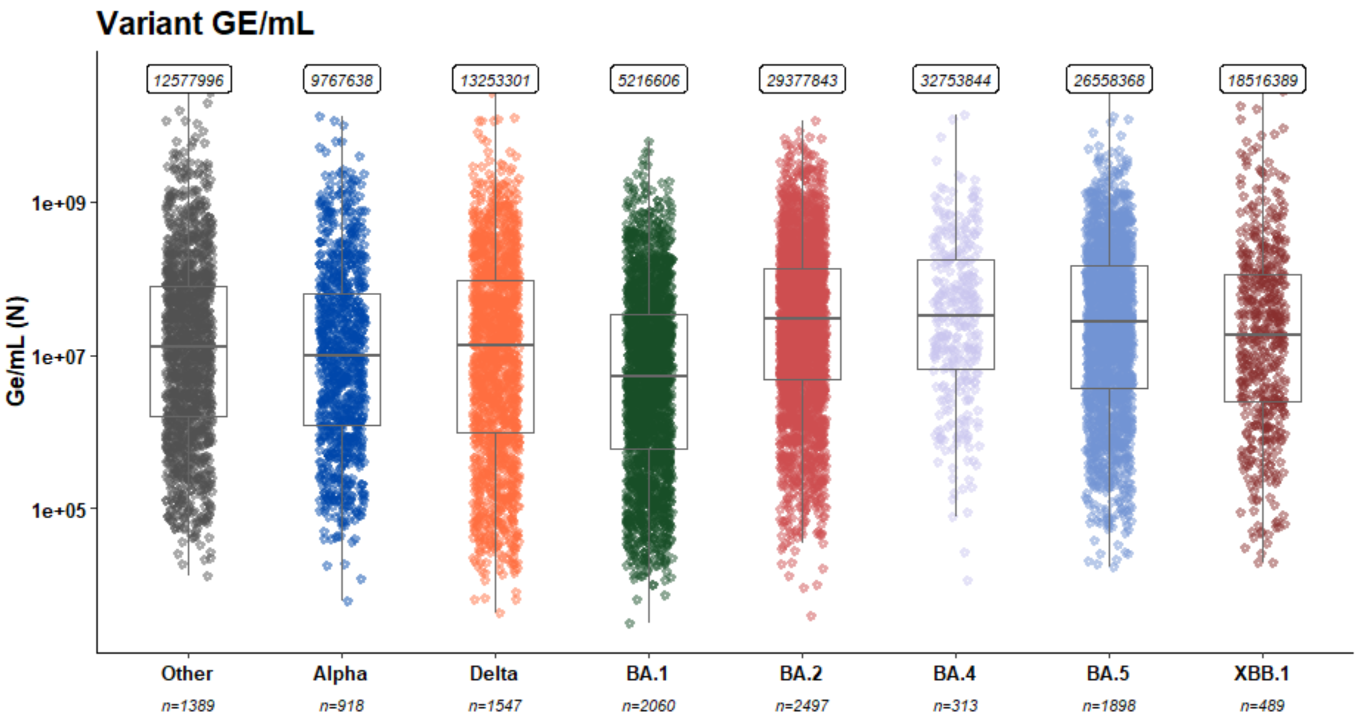


**Figure S3 Genome equivalents retrieved from qPCR assay.** qPCR Ct values were fitted to a standard curve to retrieve genome equivalents per ml.

### Supplementary Figure 4

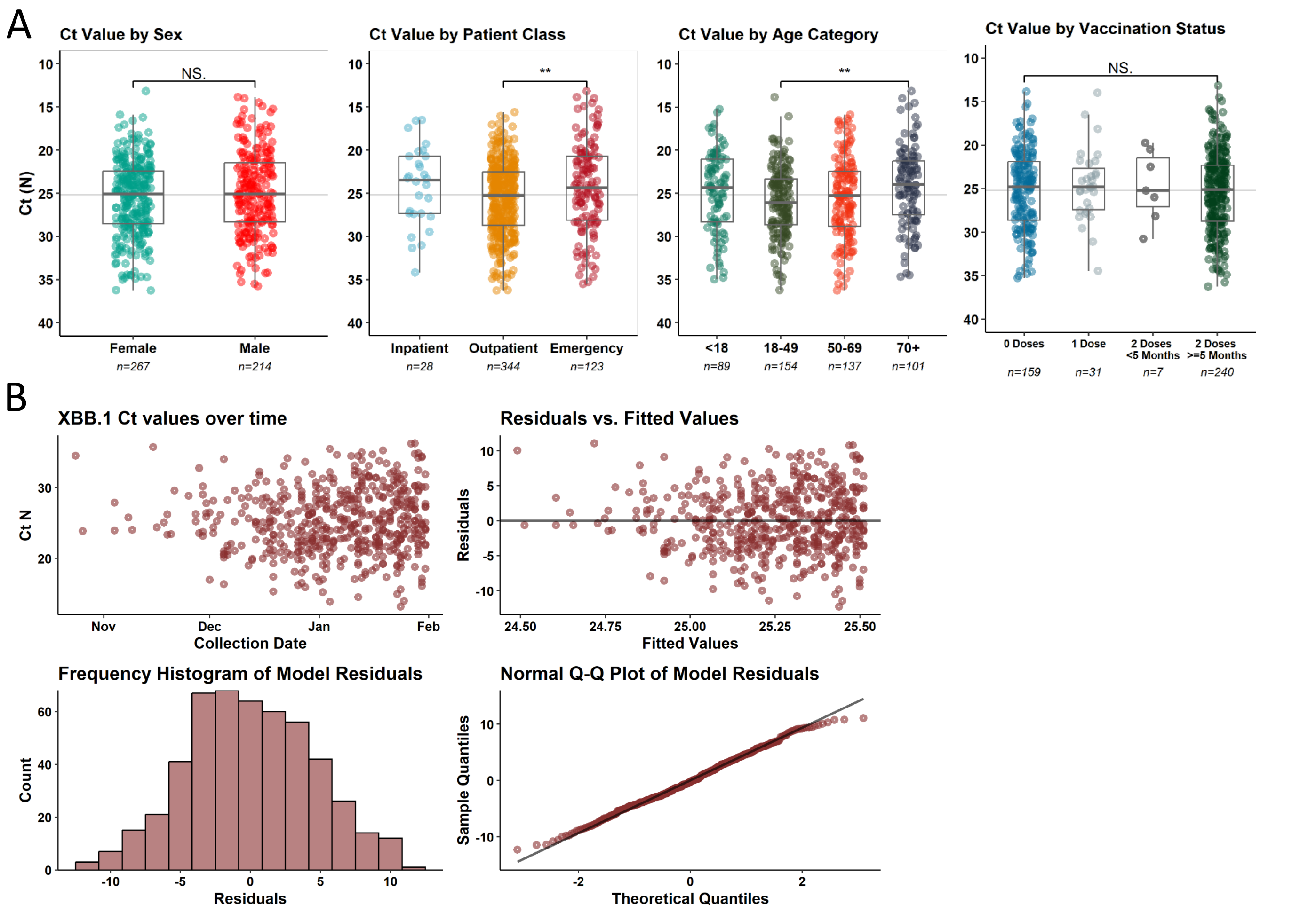


**Figure S4 Analysis of viral loads measured in early XBB samples (A)** Samples stratified according to sex, patient class, age and vaccination status **(B)** Raw data and plotted trend from XBB samples
